# Seroprevalence against COVID-19 and follow-up of suspected cases in primary health care in Spain

**DOI:** 10.1101/2020.06.13.20130575

**Authors:** C Brotons, J Serrano, D Fernández, C Garcia-Ramos, B Ichazo, J Lemaire, P Montenegro, I Moral, R Pérez Wienese, M Pitarch, M Puig, MT Vilella, J Sellarès

## Abstract

**Background:** During the coronavirus disease 2019 (COVID-19) pandemic little information has been available about patients with mild or moderate symptoms attended and followed in the primary care setting, most of whom had an unknown status for the severe acute respiratory syndrome coronavirus 2 (SARS-CoV-2) infection.

**Objectives:** We aim to measure the seroprevalence of antibodies against SARS-CoV-2 infection in a community sample of asymptomatic individuals and among symptomatic patients (without confirmed diagnosis) followed in a primary care setting.

As a secondary objective, we estimated the proportions of symptomatic patients seeing at an emergency department (ED), hospitalized or dying, and identified the most important clinical symptoms associated with a positive infection.

**Methods:** From April 21 to April 24 2020, we selected a random sample of 600 individuals stratified by age groups, from a total population of 19,899 individuals from a community area in Barcelona (study population 1). From April 29 to May 5 2020, we also invited all the patients that had been followed by general practitioners (GPs) (study population 2).

We used for both populations COVID-19 Rapid lateral flow immunoassay which qualitatively assesses the presence of patient-generated IgG and IgM in approximately 10-15 minutes.

The prevalence (95% confidence intervals [CI]) of infection (past and current) was defined as the proportion of individuals with antibody seropositivity. Odds ratios (ORs) for a positive test result were estimated using logistic regression analysis.

**Results:** Three hundred and eleven asymptomatic individuals from the randomly selected sample accepted to participate in the study. The overall mean age was 43.7 years (SD 21.79, range 1-94) and 55% were women. Seventeen individuals were seropositive for IgM and/or IgG, resulting an overall prevalence of 5,47% (95% CI, 3.44-8.58).

Six-hundred and thirty-four symptomatic patients were followed by GPs. The overall mean age was 46.97 years (SD 20.05, range 0-92) and 57.73% were women. Of these, 244 patients (38.49%) were seropositive for IgM and/or IgG.

During the follow-up period, 27.13% of symptomatic patients attended the ED, 11.83% were hospitalized and about 2% died.

Results of the multivariate logistic regression analysis showed that the OR for a positive test was significantly increased in patients who had fever (>38°C), ageusia and contact with a patient diagnosed with COVID-19.

**Conclusions:** The seroprevalence of antibodies against SARS-CoV-2 among asymptomatic individuals in the general population was lower than expected.

Approximately 40% of the symptomatic patients followed by GPs during the peak months of the pandemic in Barcelona, were positive. Fever (>38°C), anosmia, ageusia and contact with a patient diagnosed with COVID-19 were associated with a positive test result.

## Introduction

The COVID-19 pandemic is a major challenge for health systems, citizens and policy makers worldwide (1). Given the alarming levels of spread, severity of disease, and number of affected countries, on March 11th, 2020 the World Health Organization (WHO) declared COVID-19 as a pandemic (2).

A report of the London Scholl of Hygiene and Tropical medicine on April 30^th^ estimated that since Spain has recently seen a large increase in the number of deaths, and given its smaller population, 15% of the population could be infected (3).

Contrary to other countries, Spain had not implemented, until very recently, an intensive testing strategy for suspected cases of COVID-19 infection using PCR swab testing or rapid antibody testing in the primary care setting. The lack of intensive testing and subsequent tracking of cases and close contacts is likely to be one of the key factors contributing to the rapid widespread of the infection and the high number of coronavirus deaths relative to its population. Therefore, since the beginning of the pandemic we decided to implement in our primary health care area a comprehensive program which included four components: a seroprevalence study in asymptomatic patients, a follow-up study in symptomatic patients, a survey in institutionalized patients, and a survey in health care workers. Monitorization of patients reporting mild or moderate symptoms compatible with COVID-19, consisted of a close follow-up of patients by general practitioners (GPs) every 24 or 48 hours through telephone contacts to ensure that patients maintained self-isolation from day 7 to day 14 until symptoms disappeared and promptly refer patients developing severe symptoms, to the emergency department at one of the hospitals in the health care area.

We present here results of the first two components.

## Study objectives

This study aims to estimate the seroprevalence of antibodies against SARS-CoV-2 in asymptomatic individuals in a community setting, and to characterize the antibody profile in suspected cases with mild or moderate symptoms, followed by GPs since the beginning of the pandemic.

## Methods

### Study design

The seroprevalence study in asymptomatic patients used a cross-sectional survey approach to recruit individuals for participation in the study.

The follow-up study used an observational prospective follow-up approach for monitoring suspected cases of COVID-19 followed by primary care physicians.

### Study population

For the seroprevalence study (study population 1) we selected a random sample of 600 individuals one year or older, stratified by age groups (1-14,15-29,30-39,40-49,50-59,60-69,70->80 years) from a total population of 19,899 individuals registered in a primary health care center, from a community area of Barcelona, Spain. Institutionalized patients, terminally ill patients, suspected cases of COVID-19 infection, and patients tested positive for COVID-19 prior to recruitment, were excluded.

For the follow-up study (study population 2) we included all patients aged one year or older consulting the primary care physician either face-to-face or by phone with mild or moderate symptoms (without a confirmed diagnosis) during the COVID-19 pandemic from March 2 to April 24, 2020. These patients were followed mostly by phone, every 24 or 48 hours until the resolution of symptoms or referral to a hospital, as appropriate.

### Setting

Individuals from both study populations were invited to attend a municipality center for the elderly (close down during the pandemic) located near-by the primary health care center. A team of trained GPs, nurses, and medical students carried out the survey from April 21 to April 24, 2020, (study population 1) and from April 29 to May 5, 2020 (study population 2).

We used for both populations COVID-19 capillary blood draws and a lateral flow immunoassay which qualitatively assessed the presence of patient-generated IgG and IgM in about 10-15 minutes.

Data were collected through a standardized questionnaire using a web-based platform designed for the study (studies4Covid) by Universal Doctor-a social enterprise developing technology solutions for global health- and EAP Sardenya. The questionnaire was installed in tablet computers to facilitate collection of data.

All adult participants provided verbal consent to participate in the survey. For children and adolescents, written consent from a legal representative, was required.

The study design was reviewed and approved by the ethics committee of the Institut Universitari d’Investigació en Atenció Primària (IDIAP Jordi Gol) (number 20/104-P).

### Main study endpoints

The prevalence of infection (past and current) in asymptomatic individuals was defined by antibody seropositivity. For the symptomatic patients with COVID-19-compatible symptoms followed by GPs, the endpoints of interest were the proportion of patients with antibody seropositivity, the proportion of patients seeing at the emergency department (ED), the proportion of hospitalized patients and proportion of deaths.

### Statistical analysis

Descriptive statistics were calculated, using means and SD for continuous variables and frequencies and percentages with 95% CI for categorical variables.

Univariate analyses were performed including all the variables and comparing positive cases versus negative cases. Significant variables were included in a multivariate logistic regression analysis (using a stepwise procedure selection) to identify factors influencing positive antibodies test result.

All the analyses were conducted using the STATA 14 version.

## Results

### Seroprevalence study (Study population 1)

Three hundred and eleven asymptomatic individuals agreed to participate (response rate of 52%). Main reasons for non-responses included among others, difficulties in reaching people through telephone calls, concerns related to breaking of confinement and important comorbidities. The overall mean age of participants was 43.7 years (SD 21.79, range 1-94), and 55% were women.

None of the participants had been tested previously for SARS-CoV-2 by rRT-PCR.

Of the 311 participants, 17 were seropositive for IgM and/or IgG, resulting a prevalence of 5.47% (95%CI 3.44-8.58). Six patients had both IgG and IgM positive, 6 patients had only IgM positive and 5 patients has only IgG positive. Women had a higher seroprevalence than men (6.43% vs 4.28%)

Prevalence data by age groups is shown in Figure 1. The highest prevalence was found in the age group of 80 years of age or older.

**Figure 1.**
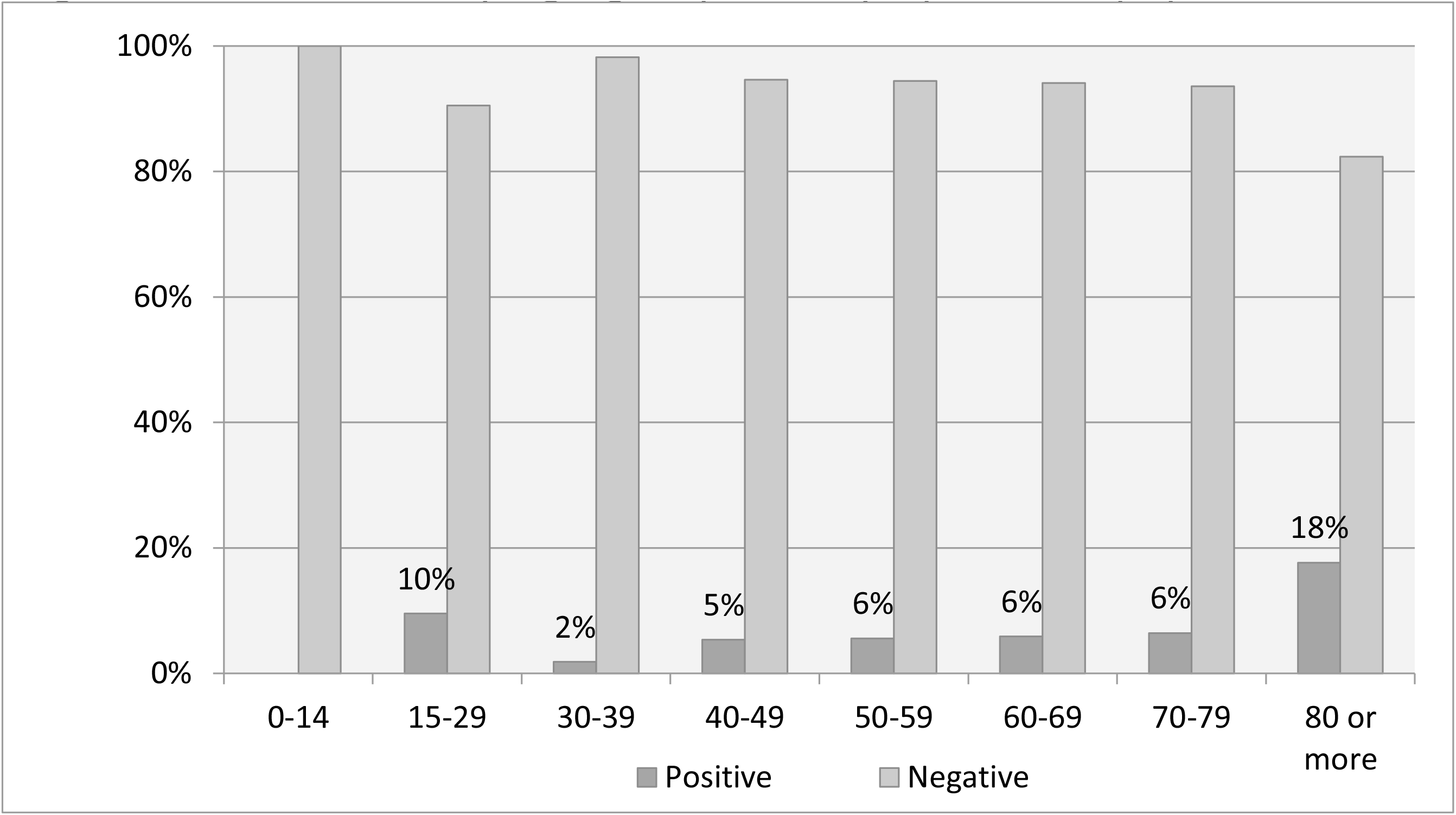
Prevalence by age groups in asymptomatic population.

### Observational study and patient follow-up (Study population 2)

During the period of follow-up, 12 patients died (4 male, 8 female), 10 of whom were seropositive.

Seven-hundred and forty-three symptomatic patients were followed by GPs for two months, and 634 underwent the serological test (participation rate of 85.33%). The overall mean age was 46.97 years (SD 20.0, range 0-92) and 57. 73% were women. Of these, 244 patients (38.49%, 95%CI 34.78%-42.33%) were seropositive for IgM and/or IgG.

During the follow-up period, 27.13% of symptomatic patients attended the ED and 11.83% were hospitalized.

Forty and one patients had a previous SARS-CoV-2 rRT-PCR performed, 25 were positive and 16 negative. Sixteen out of the 25 PCR positives were seropositive for IgG.

Characteristics of patients in the study population 2 by test result are shown in table 1.

**Table 1.** Characteristics of suspected cases by test result

|  | **Test positive**<br>**n=244**<br>**(38.49%)** | **Test negative**<br>**n=390**<br>**(61.51%)** | **Total**<br>**n=634** | **p-value** |
| --- | --- | --- | --- | --- |
| **Gender male, n (%)** | 109 (44.67%) | 159 (40.77%) | 268 (42.27%) | 0.333 |
| **Contact with positive cases, n(%)** | 124 (50.82%) | 152 (38.97%) | 276 (43.53%) | 0.003 |
| **Visit to ED, n (%)** | 88 (36.07%) | 84 (21.54%) | 172 (27.13%) | <0.001 |
| **Hospital admission, n (%)** | 59 (24.18%) | 16 (4.10%) | 75 (11.83%) | <0.001 |
| **Symptoms (last 2 months), n (%)** | 215 (88.11%) | 316 (81.03%) | 531 (83.75%) | 0.019 |
| **Nº of symptoms, mean (SD)** | 4.50 (2.99) | 3.32 (2.69) | 3.77 (2.87) | <0.001 |
| **Cough, n (%)** | 128 (52.46%) | 208 (53.33%) | 336 (53.00%) | 0.830 |
| **Tiredness, n (%)** | 144 (59.02%) | 164 (42.05%) | 308 (48.58%) | <0.001 |
| **Headache, n (%)** | 98 (40.16%) | 170 (43.59%) | 268 (42.27%) | 0.396 |
| **Fever (>38°C), n (%)** | 120 (49.18%) | 86 (22.05%) | 206 (32.49%) | <0.001 |
| **Diarrhea, n (%)** | 87 (35.66%) | 108 (27.69%) | 195 (30.76%) | 0.035 |
| **Dyspnea, n (%)** | 72 (29.51%) | 98 (25.13%) | 170 (26.81%) | 0.226 |
| **Ageusia, n (%)** | 107 (43.85%) | 60 (15.38%) | 167 (26.34%) | <0.001 |
| **Anosmia, n (%)** | 104 (42.62%) | 62 (15.90%) | 166 (26.18%) | <0.001 |
| **Sore throat, n (%)** | 51 (20.90%) | 108 (27.69%) | 159 (25.08%) | 0.055 |
| **Low grade fever (37.5°C-38°C), n (%)** | 63 (25.82%) | 82 (21.03%) | 145 (22.87%) | 0.162 |
| **Shaking chills, n (%)** | 52 (21.31%) | 72 (18.46%) | 124 (19.56%) | 0.379 |
| **Nausea, vomiting, n (%)** | 50 (20.49%) | 45 (11.54%) | 95 (14.98%) | 0.002 |
| **Skin lesions, n (%)** | 23 (9.43%) | 31 (7.95%) | 54 (8.52%) | 0.517 |

**Table 2.**
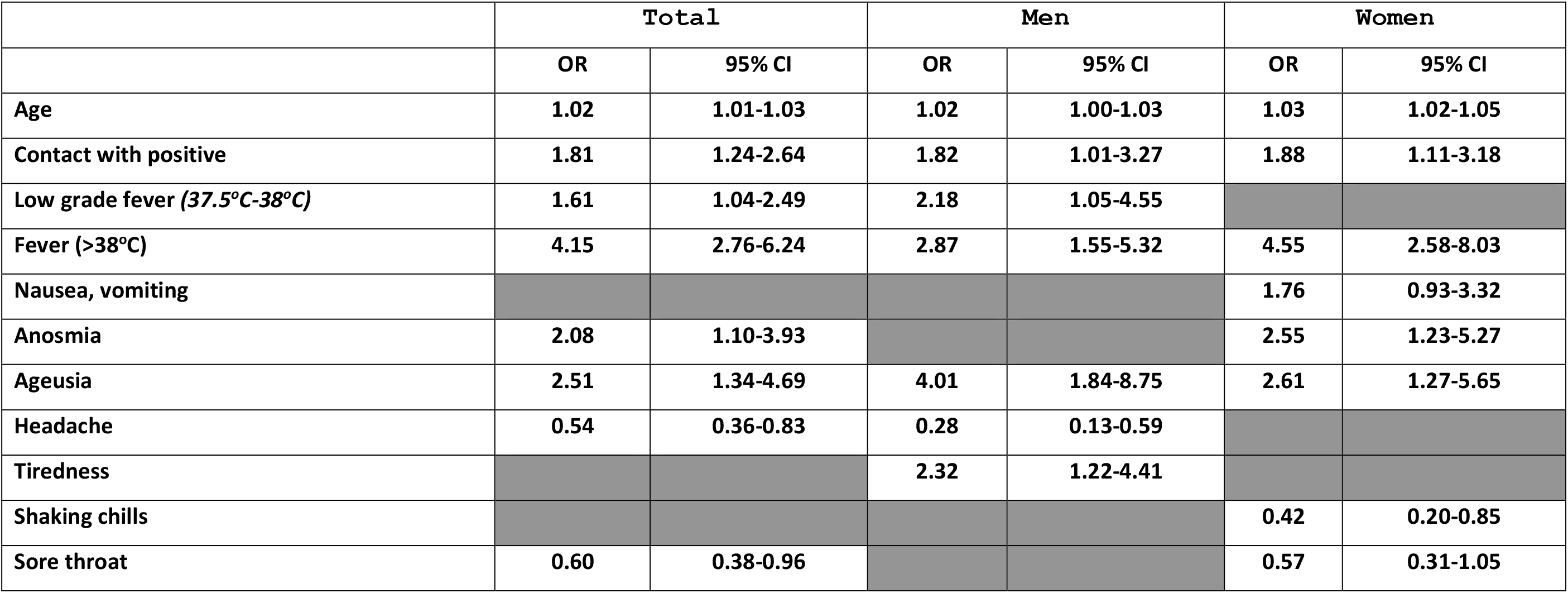
Factors associated with positive antibodies test result in symptomatic population.

Positive cases were more likely to have had close contact with other positive cases, and to have been admitted to an ED or hospitalized.

Individuals with a positive antibody positive test were more likely to have had symptoms in the last two months than individuals with a negative antibody test (88.11 vs 81.03%, p=0.019). Mean number of symptoms was higher in individuals with positive antibody test than in individuals with negative antibody test (4.50 vs 3.32, p<0.001).

The most frequent symptoms in both groups were cough, tiredness, headache and fever.

Individuals with antibody positive test were more likely to have tiredness, cough, fever, ageusia, anosmia, and headache (figure 2), with some differences between males and females.

**Figure 2.**
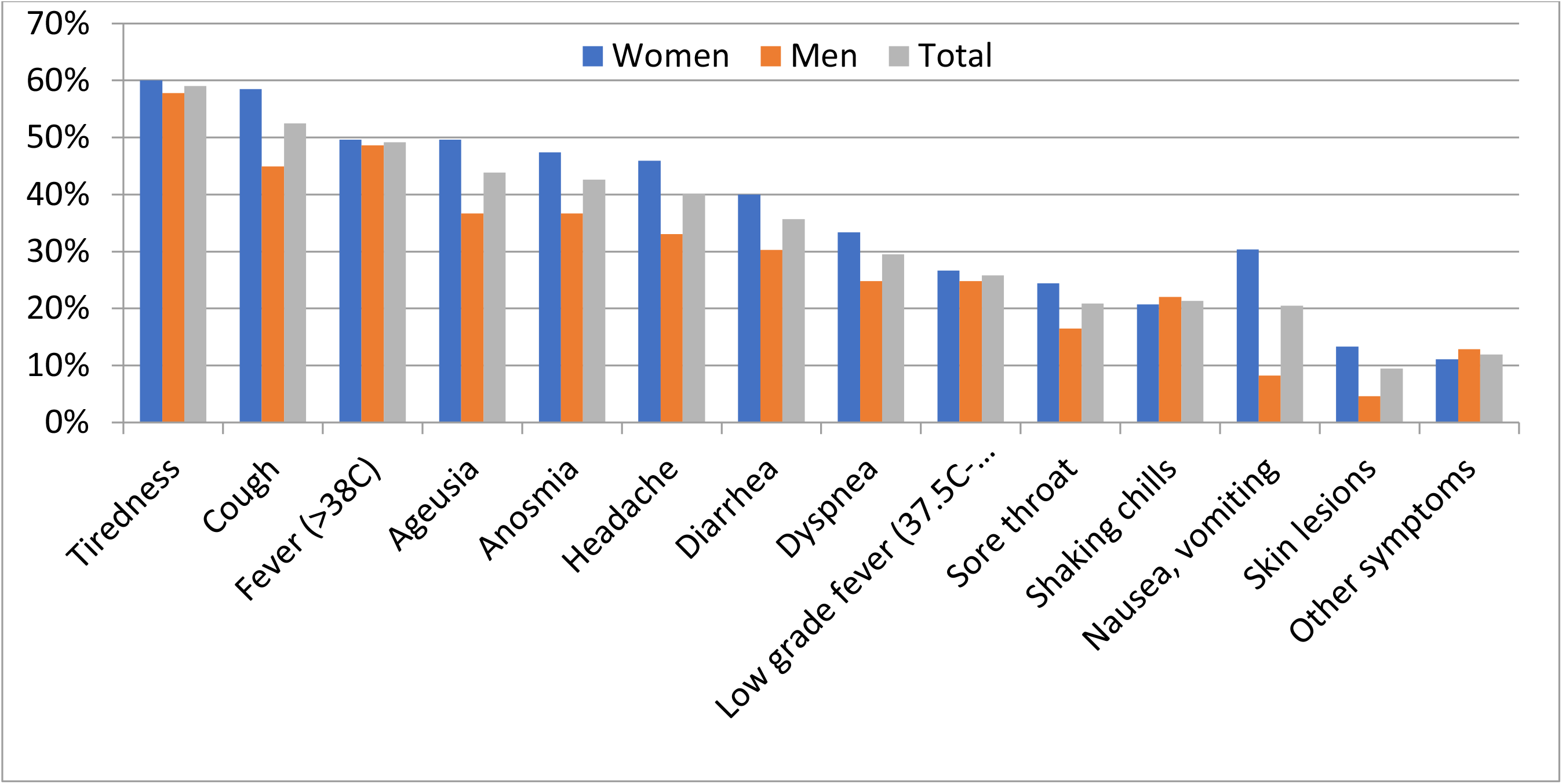
Frequency of symptoms in positive cases in symptomatic population: overall and by sex.

Results of the multivariate logistic regression analysis showed that the odds ratio (OR) for a positive test was significantly increased in patients who had fever (>38° C), ageusia and contact with a positive patient, tiredness (only men), anosmia (only in women), and decreased in patients suffering from headache (in men), sore throat (in women), and shaking chills (in women).

## Discussion

Most of the cases of COVID-19 infection are likely to be attended and managed in primary care. Therefore, effective management of COVID-19 pandemic from a primary care perspective requires estimation of the seroprevalence in a specific community area, as well as registration and follow-up of suspected or confirmed cases.

Several studies of seroprevalence have been published with varying results.

A study carried out in Santa Clara, California, in 3.330 individuals, weighting for population demographics, reported an antibody prevalence of 2.8 % (95CI 1.3-4.7%) (4).

A study conducted in Germany, among 919 individuals with evaluable infection status, 15.5% (95% CI: [12.3%; 19.0%]) were infected (5).

Researchers from Geneva, Switzerland studying a population of 1.335 individuals, observed in the first week, a seroprevalence of 3.1% (95% CI 0.2-5.99, n=343), 6.1% (95% CI 2.69.33, n=416) in the second week, and 9.7% (95% CI 6.1-13.11, n=576) in the third week (6).

In a total sample of tested company employees living in the Split-Dalmatia and Šibenik-Knin County (Croatia) antibodies were detected in 1.27% of participants (95% CI 0.77–1.98%)(7).

A study conducted on a general population sample of the island of Jersey, UK, included a total of 855 participants, showed a seroprevalence of SARS-CoV-2 antibodies of 3.1% (95% CI, 1.8– 4.4%) (8).

A study conducted in Gangelt county (N = 12 529) in Germany, comprising a sample of 1000 participants showed that 15% of the population developed antibodies to the virus [9].

Each study differs in methods used and in its sampling strategy, thus the results are not straightforward comparable.

While the results of our study are not representative for other parts of the city, they have important implications for the implementation of preventive measures to contain the COVID-19 pandemic.

Recent results from a country-wide seroprevalence study of nearly 70.000 participants in Spain, using antibody blood tests, has showed that only 5% of the population have been infected with the coronavirus (10). This result is in line with the prevalence observed in our study performed at a local level which is far below the rate that would provide the population with the so-called herd immunity, which experts place at 60% at the very least.

With regard to the frequency of symptoms we observed in our study that anosmia and ageusia are the fourth and the fifth more frequent symptoms in infected patients. Results from the logistic regression analysis showed that fever (>38^°^C), anosmia, ageusia and contact with a positive patient were risk factors associated with a positive test result. A multi-center cross-sectional cohort study in primary care patients in Germany, also showed that patients who reported anosmia had a 4-fold increase for a positive test result (OR= 4.54, 95% CI, 1.51-13.67) (11).

In a recent study done in 2153 consecutive ambulatory and hospitalized patients with positive results on reverse transcriptase polymerase chain reaction (RT-PCR) testing at 18 European hospitals found that a total of 1754 patients (87%) reported loss of smell, whereas 1136 (56%) reported taste dysfunction (12). The results of this study highlight the importance of considering loss of smell and taste in the diagnosis of mild to moderate COVID-19.

One of the limitations of our study is that characteristics of serological immunoassay tests may not be sufficiently explored and validated [13]. We did not perform RT-PCR to the IgM positive cases, thus we could not confirm if these individuals were infected. The WHO stated that serological tests, could be susceptible to cross-reaction with other frequent infections, like human coronaviruses causing common cold [14]. Nonetheless, despite their limitations, serology testing for COVID-19 are useful to better quantify the number of cases of COVID-19, including those that may be asymptomatic or have recovered (13).

## Conclusions

The seroprevalence of antibodies against SARS-CoV-2 among asymptomatic individuals in the general population was 5.47%, which was lower than expected.

Approximately 40% of the symptomatic patients followed by GPs during the peak months of the pandemic in Barcelona, were positive for antibodies against SARS-CoV-2. Thirty percent of symptomatic patients attended the ED, 13% were hospitalized and about 2% died.

Fever (>38°C), anosmia, ageusia and contact with a patient diagnosed with COVID-19 were associated with a positive test result.

## Data Availability

The authors confirm that the data supporting the findings of this study are available

## Acknowledgements

The authors wish to thank all the professionals of the Sardenya Primary Heath Care Center (EAP Sardenya), along with all study participants. We are particularly thankful to the medical students from Catalonia International University for assistance with study organization and data collection, and to employees of Universal Doctor.

## Authors contributions

CB and JS conceived and designed the study; BI, PM, MPi, MPu, MTV, DF acquired the data; CB and IM analyzed and interpreted the data; CB, IM, JS and JS drafted the manuscript; all authors critically revised the manuscript for important intellectual content; all authors approved the version to be submitted; all authors agree to be accountable for all aspects of the work.

